# Impact of Diabetes Status on Immunogenicity of Trivalent Inactivated Influenza Vaccine in Older Adults

**DOI:** 10.1101/2021.10.04.21264429

**Authors:** Sarah Spencer, Jessie R. Chung, Edward A. Belongia, Maria Sundaram, Jennifer Meece, Laura A. Coleman, Richard Zimmerman, Mary Patricia Nowalk, Krissy K. Moehling, Ted Ross, Chalise E. Carter, David Shay, Min Levine, Justine Liepkalns, Jin Hyang Kim, Suryaprakash Sambhara, Mark Thompson, Brendan Flannery

## Abstract

Individuals with type 2 diabetes mellitus experience high rates of influenza virus infection and complications. We compared the magnitude and duration of serologic response to trivalent influenza vaccine in adults aged 50-80 with and without type 2 diabetes mellitus. Serologic response to influenza vaccination was similar in both groups: greater fold-increases in antibody titer occurred among individuals with lower pre-vaccination antibody titers. Waning of antibody titers was not influenced by diabetes status.

## Background

Individuals with type 2 diabetes mellitus are at increased risk of influenza complications following influenza virus infection ^1^. Influenza vaccination has been recommended for persons of all ages with diabetes since 1960 ^2^. It is unknown if poor immunological response to vaccination contributes to the high risk of influenza-related complications among diabetic adults ^1^. Prior studies have suggested no impairment of serologic response to influenza vaccination among diabetic compared to non-diabetic adults ^3 4^. However, other factors, such as age, obesity, control of diabetes, serum vitamin D concentrations and medications may be associated with vaccine response among diabetics ^3-8^. For example, increasing age has been associated with decreased vaccine response ^5^ and obesity has been shown to be associated with increased decay of antibody titers over time ^8^. Another hypothesis suggests that immunomodulatory medications that are routinely recommended for persons with diabetes, such as statins, lead to decreased immune response to vaccination due to their anti-inflammatory effect ^9^. One study of hospitalized adults with influenza A(H1N1)pdm09 virus-associated illness found that diabetes was not associated with severity of influenza virus infection after controlling for obesity ^10^. The duration of immune response to influenza vaccination and the decline of antibody titers over time has been explored in studies of serologic response to vaccination and vaccine effectiveness ^11 12^, but has not been thoroughly investigated among diabetics. The objective of the present study was to assess whether presence of type 2 diabetes affected the magnitude and duration of antibody response to influenza vaccination among older adults.

## Methods

Participants were recruited from study sites in outpatient medical facilities in Marshfield, Wisconsin and Pittsburgh, Pennsylvania, as described previously ^13^. Approximately equal numbers of adults with type 2 diabetes and non-diabetic adults were targeted for recruitment at each site. Diabetes status was determined by medical record documentation. Eligible participants aged 50–80 years who had not received the 2011–2012 influenza vaccine were enrolled from August-November, 2011. The enrollment period preceded influenza circulation in these communities. Exclusion criteria included documented contraindications to receipt of inactivated influenza vaccine ^14^, Guillain-Barré syndrome, dementia or Alzheimer disease, estimated life expectancy <2 years, immunosuppressive medical treatment or immunocompromising condition, or concurrent participation in another influenza vaccine research study. Consented participants had blood drawn prior to receiving the 2011–2012 standard-dose trivalent inactivated influenza vaccine (IIV3) and 17-28 days post-vaccination. Participants that returned for the second year of the study received a third blood draw in the fall of 2012 prior to the circulation of influenza. Serum samples were aliquoted and frozen at –80°C until assayed. Study procedures, informed consent and data collection documents were reviewed and approved by Institutional Review Boards of Marshfield Clinic, the University of Pittsburgh, and the Centers for Disease Control and Prevention.

### Laboratory Procedures

Serology reference viruses for the 2011–2012 influenza season included A/California/07/2009 (H1N1pdm09), A/Victoria/210/2009(H3N2) (A/Perth/16/09-like), and B/Brisbane/60/2008 (Victoria lineage). In addition, sera from patients enrolled in the 2012-2013 influenza season were tested against A/California/07/2009 (H1N1pdm09), A/Victoria/361/2011(H3N2), B/Brisbane/60/2008 (Victoria lineage), and B/Wisconsin/01/2010 (Yamagata lineage) viruses. Hemagglutination inhibition (HI) assays were performed with pre- and post-vaccination serum specimens as previously described ^15^ using 0.5% turkey erythrocytes. HI assays were conducted simultaneously on paired pre- and post-vaccine sera or paired post-vaccine and day 365 sera from each participant at the Battelle Memorial Laboratory (Aberdeen, Maryland). Sera were diluted 2-fold starting from 1:10 and tested in duplicate. The HI titer was the reciprocal of the serum dilution in the last well with complete hemagglutination inhibition. The final HI titer was estimated as the geometric mean of duplicate samples; a value of 5 was used for HI <10.

Induction of vaccine antigen-specific memory B-cells (IgG, IgM and IgA) was evaluated by an ELISPOT assay, using a paired set of day 0 and day 21 peripheral blood mononuclear cells (PBMCs) stimulated *in vitro* for 5 days with polyclonal stimuli as previously described ^16^. For serum vitamin D levels, 25-hydroxyvitamin D concentrations (ng/mL) were measured with a Waters ultra-performance liquid chromatography with tandem mass spectrometer, as previously described ^6^.

### Statistical Analysis

We compared descriptive characteristics of diabetic and non-diabetic participants using the χ ^2^ test for categorical variables and Student’s *t*-test for continuous variables. Geometric mean titers (GMT), GMT ratios, and 95% confidence intervals (CI) were calculated using repeated measures linear mixed models as previously described ^16^. Seroconversion was defined as a four-fold rise or greater in HI titer with a final titer ≥40. Seroprotection was defined as titer ≥40 on the second serum sample. Rate of change between Day 21 and Day 365 was defined as the difference in log_2_-transformed titer. Time in days to decrease one 2-fold dilution in HI titer was calculated as the reciprocal of the model estimated rates, assuming linear (log_2_) decay over time, as described ^11^. We used linear regression with log_2_-transformed fold-rise as the dependent variable to identify associations between antibody waning and factors including pre-vaccination HI titer, post-vaccination titer, age (in years), diabetes status, serum vitamin D concentration (<30 or ≥30 ng/mL) ^6^, and impaired functional status (positive response to any of five functional status-related questions [Supplemental Table 2]). Those who seroconverted between Day 21 and Day 365 were excluded from analyses of Day 365. Predictors of the fold-rise between Day 0 and Day 21 and predictors of the rate of change between Day 21 and Day 365 were examined using linear regression models. Induction of memory B-cell responses was summarized as geometric mean percentages (GMP) ratio following estimation of means and differences in means of GMPs at days 0 and 21, as previously described ^16^. All analyses were conducted using SAS statistical software (version 9.3; SAS Institute, Inc., Cary, NC).

## Results

A total of 92 participants with type 2 diabetes (70 in Wisconsin and 22 in Pennsylvania [Supplemental Table 1]) and 113 non-diabetic individuals (80 in Wisconsin and 33 in Pennsylvania) were enrolled before the 2011–2012 influenza season; proportions of enrollees with diabetes were similar at both enrollment sites (Supplemental Table 1). Diabetic enrollees were more likely to be male (p=0.03) and were older (p=0.001) than non-diabetic enrollees. Diabetics enrollees also had higher BMI (p<0.001), were more likely to be obese (BMI ≥30, p<0.001), and had lower self-rated general health status (p<0.001). Among 88 diabetics for whom hemoglobin A1c was available, 48 (55%) had controlled diabetes (HbA1c ≤7.0%) (data not shown) ^17^. There were no differences between diabetic and non-diabetic adults in self-rated functional status measures (Supplemental Table 2). A total of 190 (93%) participants (84 [91%] diabetic patients and 106 [94%] non-diabetic patients) returned before the 2012–2013 influenza season and provided a day 365 (D365) serum specimen.

Pre-vaccination (D0), post-vaccination (D21) and D365 HI titers were measured for three 2011–2012 serology reference viruses (A/California/07/2009 [H1N1pdm09], A/Victoria/210/2009 [H3N2], and B/Brisbane/60/2008 [Victoria]) and two 2012–2013 reference viruses (A/Victoria/361/2011 [H3N2] and B/Wisconsin/01/2010 [Yamagata]) (Figure). Baseline titer and percent seroprotection at D0 and D365 did not differ by diabetes status (Table). Percent seroprotection at day 21 (D21) was significantly higher for non-diabetics for influenza A(H3N2) viruses but was similar for influenza A(H1N1)pdm09 and influenza B viruses. Pre- and post-vaccination GMT ratios (i.e., D21/D0) did not differ significantly by diabetes status. From the linear regression model, only pre-vaccination titer was significantly associated (negatively) with D21/D0 GMT ratio when controlling for age and diabetes status; participants with higher pre-vaccination titers had lower D21/D0 GMT ratios for all antigens tested (data not shown). Consistent with GMT ratios, induction of vaccine-specific memory B cells (IgG, IgM and IgA) at D21 was comparable between diabetic and non-diabetic persons (Supplemental Table 4).

**Figure.**
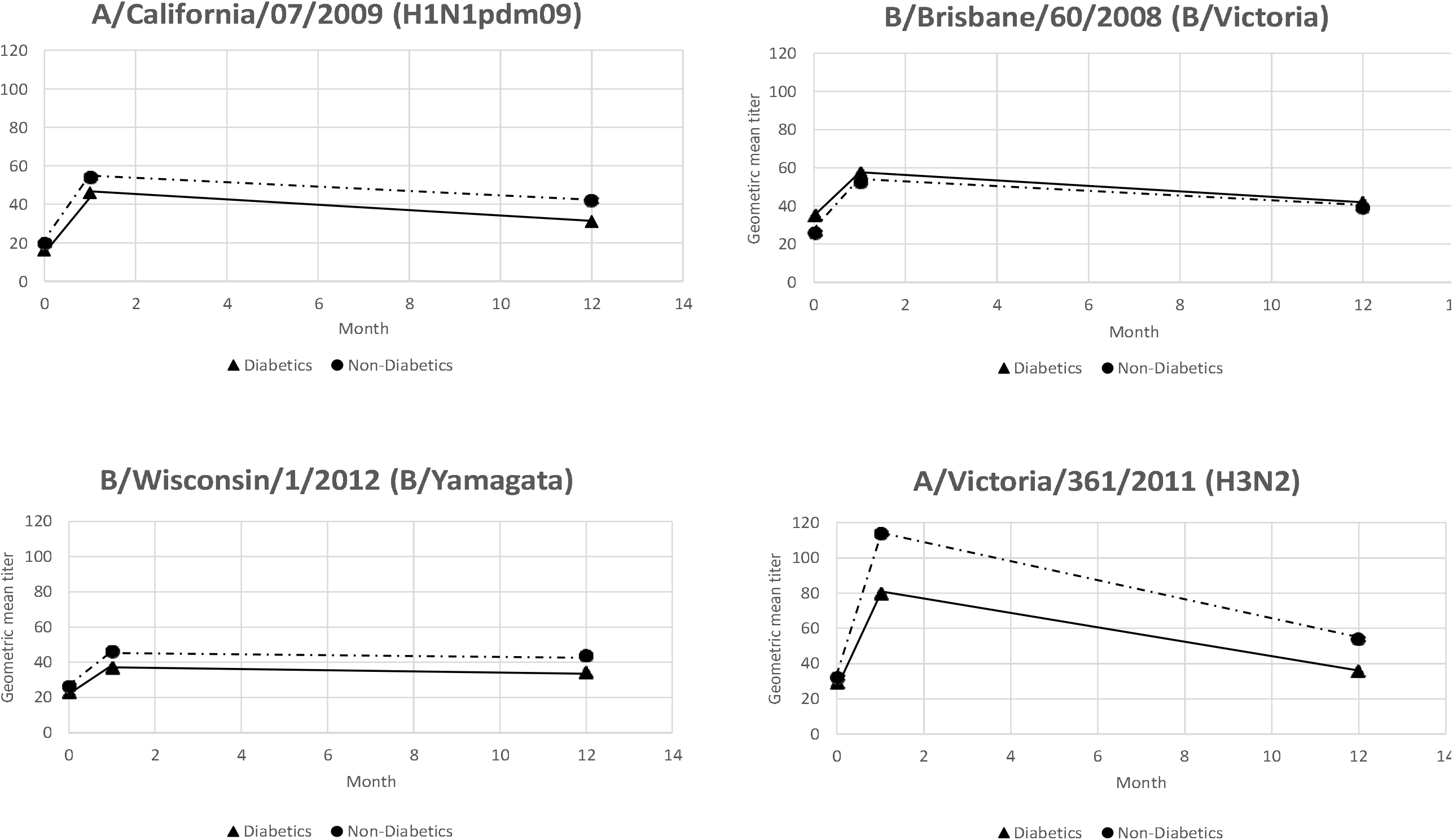
Pre- and post-vaccination^1^ hemagglutination inhibition (HI) titers to influenza vaccine reference antigens among individuals aged 50-80 years with and without type 2 diabetes mellitus^2^. ^1^D0 pre-vaccination, D21 post-vaccination, and D326 post-vaccination ^2^Figure represents linear approximations

**Table.**
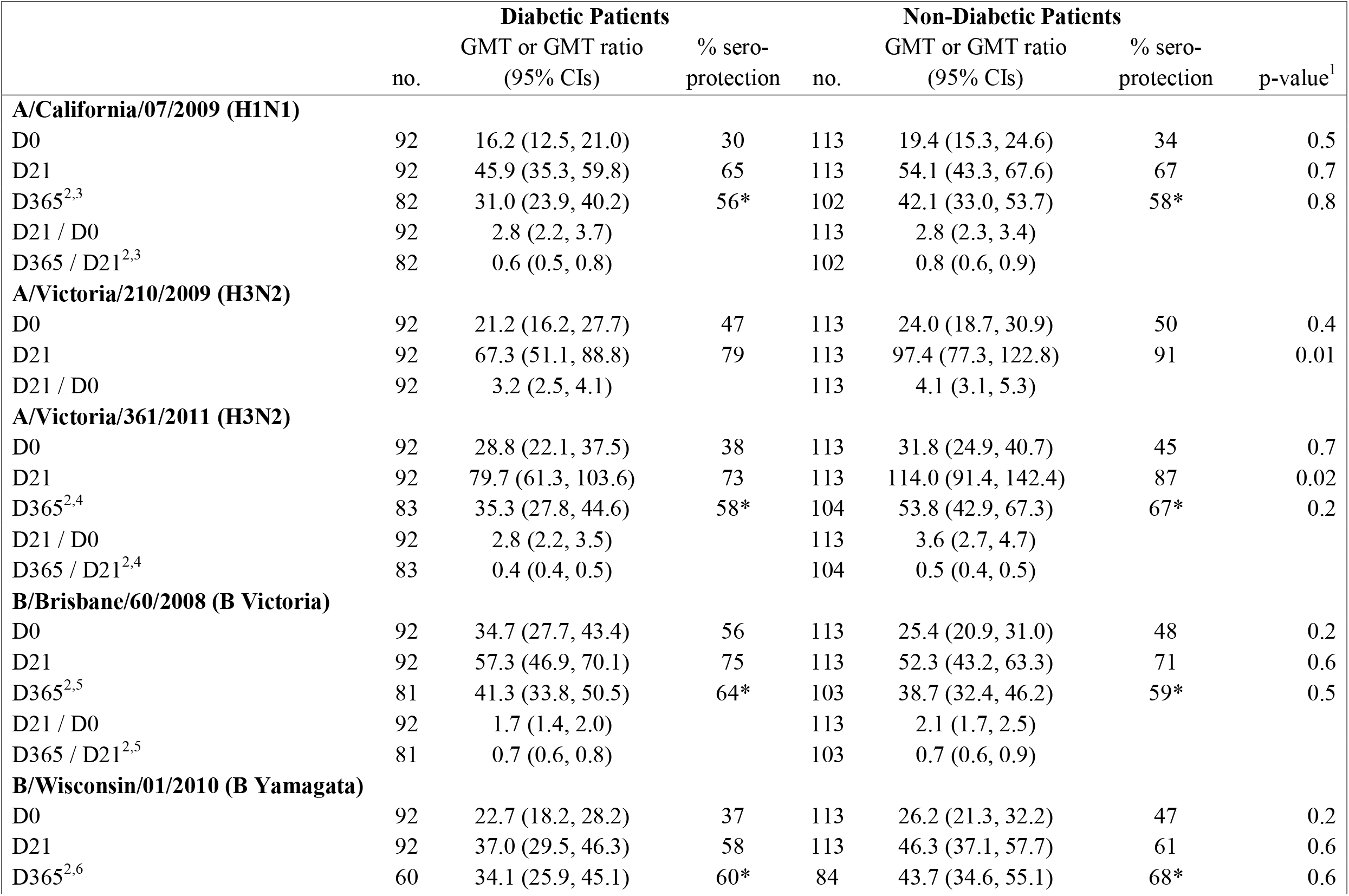
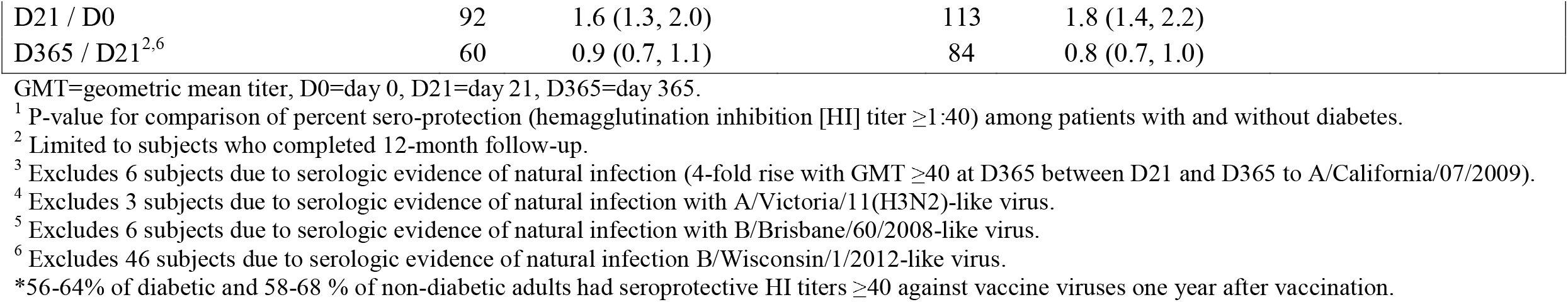
Pre- and post-vaccination hemagglutination inhibition (HI) titers to influenza vaccine reference antigens among individuals aged 50-80 years with and without type 2 diabetes mellitus.

HI titers were measured for four reference viruses for participants who returned for the D365 specimen collection (Figure). D365/D21 GMT ratios did not differ by diabetes status for any of the antigens measured (p>0.05); 56-64% of diabetic and 58-68% of non-diabetic adults had seroprotective HI titers ≥40 against vaccine viruses one year after vaccination (Table, D365 values). Furthermore, diabetes was not a significant predictor in the linear regression model where the rate of decline in antibody titer was the outcome for the antigens measured (Supplemental Table 3). Extrapolating from antibody declines from D21 to D365, assuming linear trends, the estimated number of days until post-vaccination HI titers decreased 2-fold would be 675 days for A/California/07/2009, 717 days for B/Brisbane/60/2008, 1075 for B/Wisconsin/01/2010 and 294 for the 2012-2013 H3N2 virus A/Victoria/361/2011. From the linear regression model controlling for age and diabetes status, antibody titer at D21 was the only significant predictor of the rate of antibody decline (Supplemental Table 3). Participants with higher D21 post-vaccination titers had steeper decline over the 12-month period. Diabetes, age, vitamin D level, and BMI were not significant predictors for any of the viruses tested.

## Discussion

In this study, adults aged 50-80 years with and without type 2 diabetes mellitus exhibited similar serologic response to influenza vaccination and persistence of elevated antibody titers. Both diabetic and non-diabetic adults responded to inactivated influenza vaccine. Seroconversion was similar against all vaccine components, as well as seroprotection against A(H1N1)pdm09 and B virus vaccine components, while seroprotection against the influenza A(H3N2) virus vaccine component was higher among adults without diabetes compared to non-diabetic adults. Diabetes was not significantly associated with antibody response or induction of memory B cells to vaccine components in models controlling for age, obesity and other potential predictors of response. Pre-vaccination HI titer was the strongest predictor of post-vaccination (D21) titer, with lower pre-vaccination HI titers associated with greater fold-rise in D21/D0 GMT ratio. These results are consistent with previous serologic studies that showed no impairment of initial immune responses to vaccine among adults with diabetes ^3 4^. Declines in antibody titers were also similar among diabetic and non-diabetic adults; excluding subjects with serologic evidence of infection during the 2011-2012 season (Table, D365 values).

Among patients enrolled in this study, we observed no differences in the relationship between diabetes status and HI titer by subject age. One study found improved immune response to influenza vaccination among diabetics compared to non-diabetic older adults (aged ≥65 years), while no difference was observed among immune response in younger adults ^18^.

The current study provides more detail about the magnitude and duration of antibody responses to influenza vaccine in a well-characterized group of older adults than previous studies by assessing changes in GMTs rather than only seroconversion and seroprotection. One important finding of this study was the rates of waning of antibody titers among diabetic adults, which were similar to those reported among healthy adults who received inactivated influenza vaccine in a clinical trial which found that HI titers decreased slowly over 18 months ^11^.

These results are subject to several limitations. This study only included individuals with type 2 diabetes and may not be applicable to influenza vaccine response and duration of response in individuals with type 1 diabetes ^7^. Enrollees may differ from non-participants in their level of control of diabetes, prevalence of comorbidities, or behaviors (such as smoking) associated with immune response. Furthermore, no surveillance was conducted among enrolled individuals to identify influenza infections during 2011-12 season which may have contributed to a rise in HI titer; not all patients with increased HI titers excluded from analysis of duration of antibody response may have had influenza infection. Additionally, we were unable to evaluate whether or not medications influenced serologic response. Finally, HI titers are imprecise correlates of protection; we use HI titers ≥40 as a widely accepted correlate of 50% protection against influenza illness among adults ^19^.

In conclusion, diabetic and non-diabetic adults appear to respond similarly to influenza vaccination and retain elevated antibody levels until a subsequent season. Efforts should continue to increase influenza vaccination coverage among all adults, especially those at increased risk of severe disease, such as people with diabetes.

## Data Availability

Data are not publicly available at this time.

