## Supplemental Tables for "Impact of Diabetes Status on Immunogenicity of Trivalent Inactivated Influenza Vaccine in Older Adults"

**Supplemental Table 1. Descriptive characteristics of diabetic and non-diabetic participants**

|  | **Diabetic (N=92)** | | **Non-Diabetic (N=113)** | | **p-value** |
| --- | --- | --- | --- | --- | --- |
|  | **N (%) or mean (sd)** | | **N (%) or mean (sd)** | |  |
| **Age, Mean** | 65.6 | (7.7) | 61.9 | (7.4) | 0.001 |
| **Sex** |  |  |  |  | 0.03 |
| Male | 49 | (53) | 43 | (38) |  |
| Female | 43 | (47) | 70 | (62) |  |
| **Site** |  |  |  |  | 0.4 |
| Wisconsin | 70 | (76) | 80 | (71) |  |
| Pennsylvania | 22 | (24) | 33 | (29) |  |
| **Race/Ethnicity** |  |  |  |  | 0.7 |
| White, non-Hispanic | 78 | (85) | 100 | (89) |  |
| Black, non-Hispanic | 13 | (14) | 11 | (10) |  |
| Other | 1 | (1) | 2 | (2) |  |
| **Body Mass Index (kg/m^2^)** | 35.2 | (7.4) | 30.3 | (6.5) | <0.001 |
| **Obese (Body Mass Index ≥30 kg/m^2^)** | 69 | (75) | 55 | (49) | <0.001 |
| **HbA1c^1^ (n=88)** | 7.3 | (1.3) | NA |  | NA |
| **Vitamin D level (ng/mL)** | 43.8 | (17.0) | 41.2 | (16.9) | 0.28 |
| **Vitamin D <30 ng/mL** | 17 | (18.5) | 25 | (22.1) | 0.52 |
| **Smoked >100 cigarettes in lifetime** | 51 | (55) | 49 | (44) | 0.1 |
| **Current health assessment^2^**  **Scale 0 (worst) – 100 (best)** | 68.7 | (17.3) | 81.4 | (13.3) | <0.001 |

^1^ Serum HbA1c concentration was measured for 88 subjects with diabetes.

^2^ The validated EQ-5D health ruler [20] was used to assess patient health on the day of enrollment. Data are presented on a scale of 1 (worst) to 100 (best).

**Supplemental Table 2. Frequency of responses to functional status measures among diabetic and non-diabetic adult participants.**

|  | **Diabetic (N=92)** | **Non-Diabetic (N=113)** | **p value** |
| --- | --- | --- | --- |
|  | **N (%)** | **N (%)** |  |
| **Self-reported mobility** |  |  | 0.5 |
| No problems walking about | 47 (51) | 66 (58) |  |
| Some problems walking about or confined to bed | 45 (49) | 47 (42) |  |
| **Self-rated ability to perform self-care** |  |  | 0.7 |
| No problems | 65 (71) | 80 (71) |  |
| Some problems washing/dressing or unable to  wash/dress self | 27 (29) | 33 (29) |  |
| **Usual activities** |  |  | 0.4 |
| No problems performing usual activities | 53 (58) | 75 (66) |  |
| Some problems performing usual activities or  unable to perform usual activities | 38 (42) | 38 (34) |  |
| **Pain** |  |  | 0.1 |
| I have no pain or discomfort | 30 (33) | 53 (47) |  |
| Moderate or extreme pain or discomfort | 62 (67) | 60 (53) |  |
| **Anxiety** |  |  | 0.8 |
| Not anxious/depressed | 60 (65) | 77 (68) |  |
| Moderately/extremely depressed | 32 (35) | 36 (32) |  |

**Supplemental Table 3. Predictors of rate of change between D21 and D365**

|  | **A/CA/07/09(H1N1)pdm09** | | **A/Victoria/361/11(H3N2)** | | **B/Brisbane/60/08 (Vic)** | | **B/Wisconsin/1/10(Yam)** | |
| --- | --- | --- | --- | --- | --- | --- | --- | --- |
|  | Coefficient (SE) | p-value | Coefficient (SE) | p-value | Coefficient (SE) | p-value | Coefficient (SE) | p-value |
| Age | -0.02 (0.01) | 0.18 | -0.02 (0.01) | 0.18 | 0.02 (0.01) | 0.05 | -0.01 (0.01) | 0.67 |
| Diabetes | -0.25 (0.20) | 0.23 | -0.15 (0.18) | 0.43 | -0.04 (0.16) | 0.79 | -0.001 (0.23) | 0.99 |
| White race | 0.29 (0.30) | 0.33 | -0.41 (0.28) | 0.14 | -0.04 (0.23) | 0.87 | 0.29 (0.31) | 0.36 |
| Sex | 0.01 (0.19) | 0.98 | -0.22 (0.17) | 0.21 | -0.14 (0.15) | 0.35 | -0.06 (0.21) | 0.78 |
| Body mass index | 0.02 (0.01) | 0.14 | -0.02 (0.01) | 0.23 | -0.01 (0.01) | 0.41 | -0.02 (0.02) | 0.26 |
| Impaired functional status^1^ | -0.39 (0.20) | 0.06 | -0.05 (0.18) | 0.77 | -0.29 (0.16) | 0.08 | -0.12 (0.24) | 0.63 |
| Vitamin D level | 0.32 (0.24) | 0.20 | 0.23 (0.22) | 0.30 | 0.15 (0.19) | 0.42 | 0.37 (0.27) | 0.17 |
| D21 HI titer | -0.27 (0.06) | <0.01 | -0.27 (0.05) | <0.01 | -0.36 (0.05) | <0.01 | -0.38 (0.06) | <0.01 |

Note: Separate linear regression models were estimated for HI titers to each reference virus.

^1^ A person was considered to have impaired functional status if he/she indicated any problems with mobility, ability to perform self-care, usual activities, pain, or anxiety.

**Supplemental Table 4. Pre- and post-vaccination frequency of antibody-secreting cells (ASCs) to influenza vaccine reference antigens among individuals aged 50-80 years with and without type 2 diabetes mellitus.**

|  | **Diabetic (N=9)** | | | | **Non-Diabetic (N=14)** | | |  |
| --- | --- | --- | --- | --- | --- | --- | --- | --- |
|  | GMP (CIs) | | GMP ratio  (D21/D0) | | GMP (CIs) | | GMP ratio (D21/D0) | P value^1^ |
|  | D0 | D21 |  |  | D0 | D21 |  |  |
| **A/CA/07/09 (H1N1)pdm09** | | | | | | | |  |
| IgG | 0.4 (0.12,1.36) | 3.58 (1.93, 6.62) | | 8.88 (2.66, 29.65) | 0.81 (0.33, 2.03) | 4.36 (2.42, 7.86) | 5.37 (1.92, 15.06) | NS |
| IgM | 2.2 (0.38, 12.74) | 10.04 (4.39, 22.98) | | 4.57 (1.22, 17.09) | 7.36 (2.81, 19.28) | 7.32 (3.92, 13.69) | 0.99 (0.59, 1.68) | 0.01 |
| IgA | 0.05 (0.01, 0.18) | 0.5 (0.12, 2.03) | | 10.37 (1.62, 66.42) | 0.21 (0.07, 0.62) | 0.77 (0.22, 2.72) | 3.73 (0.72, 19.29) | NS |
| **A/Victoria/210/09 (H3N2)** | | | | | | | |  |
| IgG | 0.12 (0.03, 0.55) | 2.43 (1.07, 5.54) | | 20.31 (2.54, 162.17) | 0.29 (0.1, 0.86) | 1.92 (0.49, 7.5) | 6.66 (2.32, 19.1) | NS |
| IgM | 9.38 (3.35, 26.32) | 10.43 (4.64, 23.45) | | 1.11 (0.44, 2.8) | 9.68 (3.42, 27.4) | 9.91 (4.51, 21.8) | 1.02 (0.54, 1.93) | NS |
| IgA | 0.28 (0.05, 1.63) | 0.57 (0.15, 2.18) | | 2.07 (0.23, 18.26) | 0.28 (0.09, 0.87) | 0.43 (0.09, 1.95) | 1.54 (0.64, 3.72) | NS |
| **B/Brisbane/60/2008 (Victoria)** | | | | | | | |  |
| IgG | 0.2 (0.05, 0.81) | 2.81 (1.52, 5.2) | | 14.17 (2.92, 68.81) | 0.82 (0.34, 1.96) | 2.88 (1.04, 7.96) | 3.5 (1.16, 10.59) | NS |
| IgM | 0.78 (3.3, 18.66) | 8.65 (4.26, 17.53) | | 1.1 (0.62, 1.95) | 7.26 (3.08, 17.15) | 9 (3.71, 21.79) | 1.24 (0.54, 2.82) | NS |
| IgA | 0.26 (0.04, 1.83) | 1.38 (0.26, 7.33) | | 5.33 (0.76, 37.55) | 0.69 (0.31, 1.55) | 1.35 (0.51, 3.57) | 1.96 (0.86, 4.47) | NS |

GMP=geometric mean percentage; NS=non-significant (p>0.05).

^1^ Comparison of GMP ratio (D21/D0) between diabetic and non-diabetic groups.
